# A Saudi G6PD Deficient Girl Died with Pediatric Multisystem Inflammatory Syndrome-COVID-19

**DOI:** 10.1101/2020.07.08.20137497

**Authors:** Maryam A. Al-Aamri, Fatimah T. Al-Khars, Sami J. Alkhwaitem, Abdulaziz K. AlHassan, Ali M. Al Aithan, Fatima H. Alkhalifa, Sameer Y. Al-Abdi

## Abstract

Reports on pediatric multisystem inflammatory syndrome (PMIS) temporally related to coronavirus disease 2019 (COVID-19) are increasing. African and Afro-Caribbean children constituted about 37% of the cases in these reports. Although glucose-6-phosphatase dehydrogenase (G6PD) deficiency is common among this population, the G6PD status of these cases has not been reported. We report the first case of PMIS related to COVID-19 from Saudi Arabia. This case was a Saudi G6PD deficient girl who died with PMIS related to COVID-19. G6PD deficiency induces redox imbalance and exaggerates the inflammatory response; thus, it might contribute to the development or the grave outcome of our case. We urgently need to assess the association between G6PD deficiency and COVID-19 in a large study as the G6PD deficiency may be a useful predictor for the progression of the COVID-19.

## Introduction

Reports on pediatric multisystem inflammatory syndrome (PMIS), also known as a multisystem inflammatory syndrome in children, temporally related to coronavirus disease 2019 (COVID-19) are increasing.^1-13^ Data on race or ethnicity are available for 392 children with probable, possible, or confirmed PMIS related to COVID-19 in nine studies form Europe and USA.^1-9^ Pooled data show that the African and Afro-Caribbean or black children constituted 37% (144/392) of these cases. Four out of the five dead children whose race or ethnicity have been reported were black.^1, 2^ Although glucose-6-phosphatase dehydrogenase (G6PD) deficiency is common among this population,^14^ the G6PD status of these cases has not been reported in these reports. We report the first case of PMIS related to COVID-19 from Saudi Arabia. This case was a previously healthy Saudi G6PD deficient girl.

### Case Report

This case is a 10–15 years old previously healthy Saudi girl. She underwent a nasopharyngeal swab for acute respiratory syndrome coronavirus 2 (SARS-CoV-2) during a COVID-19 contact tracing process. Her reverse transcription-polymerase chain reaction (RT-PCR) reported positive for SARS-CoV-2. She was asymptomatic for 22 days, and then she started to have a fever, abdominal pain, vomiting, and diarrhea. Two days later, she presented to our emergency room with a clinical picture similar to Kawasaki disease shock syndrome. She rapidly developed multiple organ dysfunction syndrome and succumbed to death despite aggressive medical support. Figure 1 and Table 1 summarize timeline events, investigations, and management of the case.

**Table 1.**
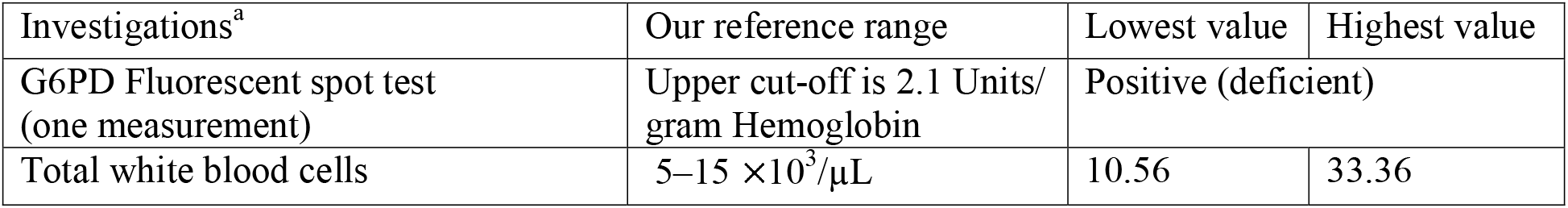
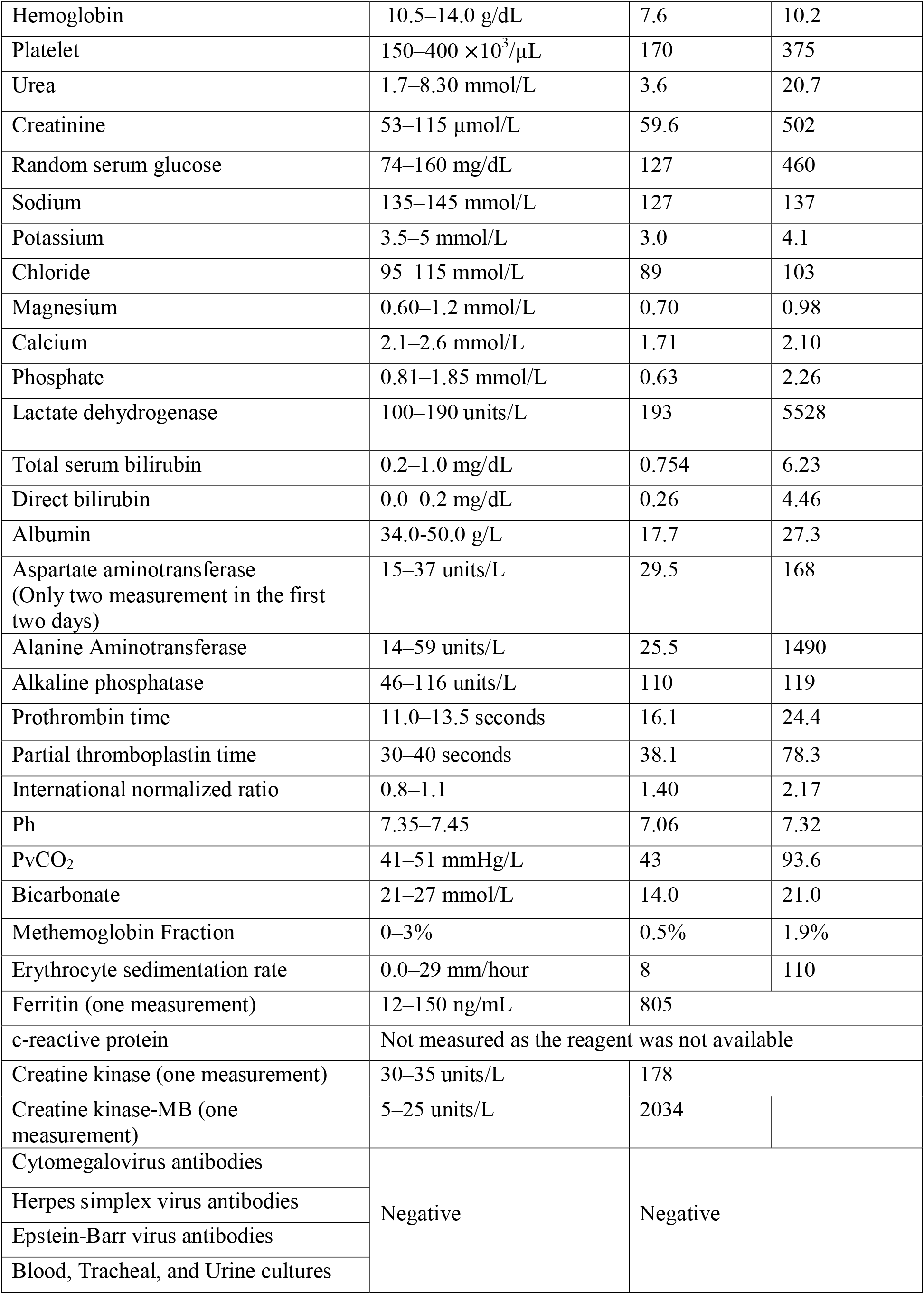
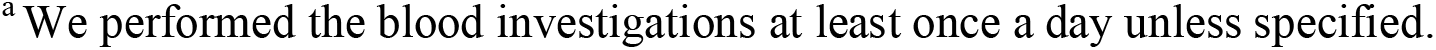
Results of Blood investigations.

| Investigations<sup>a</sup> | Our reference range | Lowest value | Highest value |
| --- | --- | --- | --- |
| G6PD Fluorescent spot test (one measurement) | Upper cut-off is 2.1 Units/gram Hemoglobin | Positive (deficient) |  |
| Total white blood cells | 5–15 × 10<sup>3</sup>/μL | 10.56 | 33.36 |
| Hemoglobin | 10.5–14.0 g/dL | 7.6 | 10.2 |
| Platelet | 150–400 ×10<sup>3</sup>/μL | 170 | 375 |
| Urea | 1.7–8.30 mmol/L | 3.6 | 20.7 |
| Creatinine | 53–115 μmol/L | 59.6 | 502 |
| Random serum glucose | 74–160 mg/dL | 127 | 460 |
| Sodium | 135–145 mmol/L | 127 | 137 |
| Potassium | 3.5–5 mmol/L | 3.0 | 4.1 |
| Chloride | 95–115 mmol/L | 89 | 103 |
| Magnesium | 0.60–1.2 mmol/L | 0.70 | 0.98 |
| Calcium | 2.1–2.6 mmol/L | 1.71 | 2.10 |
| Phosphate | 0.81–1.85 mmol/L | 0.63 | 2.26 |
| Lactate dehydrogenase | 100–190 units/L | 193 | 5528 |
| Total serum bilirubin | 0.2–1.0 mg/dL | 0.754 | 6.23 |
| Direct bilirubin | 0.0–0.2 mg/dL | 0.26 | 4.46 |
| Albumin | 34.0-50.0 g/L | 17.7 | 27.3 |
| Aspartate aminotransferase<br>(Only two measurement in the first two days) | 15–37 units/L | 29.5 | 168 |
| Alanine Aminotransferase | 14–59 units/L | 25.5 | 1490 |
| Alkaline phosphatase | 46–116 units/L | 110 | 119 |
| Prothrombin time | 11.0–13.5 seconds | 16.1 | 24.4 |
| Partial thromboplastin time | 30–40 seconds | 38.1 | 78.3 |
| International normalized ratio | 0.8–1.1 | 1.40 | 2.17 |
| Ph | 7.35–7.45 | 7.06 | 7.32 |
| PvCO<sub>2</sub> | 41–51 mmHg/L | 43 | 93.6 |
| Bicarbonate | 21–27 mmol/L | 14.0 | 21.0 |
| Methemoglobin Fraction | 0–3% | 0.5% | 1.9% |
| Erythrocyte sedimentation rate | 0.0–29 mm/hour | 8 | 110 |
| Ferritin (one measurement) | 12–150 ng/mL | 805 |  |
| c-reactive protein | Not measured as the reagent was not available |  |  |
| Creatine kinase (one measurement) | 30–35 units/L | 178 |  |
| Creatine kinase-MB (one measurement) | 5–25 units/L | 2034 |  |
| Cytomegalovirus antibodies | Negative | Negative |  |
| Herpes simplex virus antibodies |  |  |  |
| Epstein-Barr virus antibodies |  |  |  |
| Blood, Tracheal, and Urine cultures |  |  |  |

<sup>a</sup>We performed the blood investigations at least once a day unless specified.

**Figure 1.**
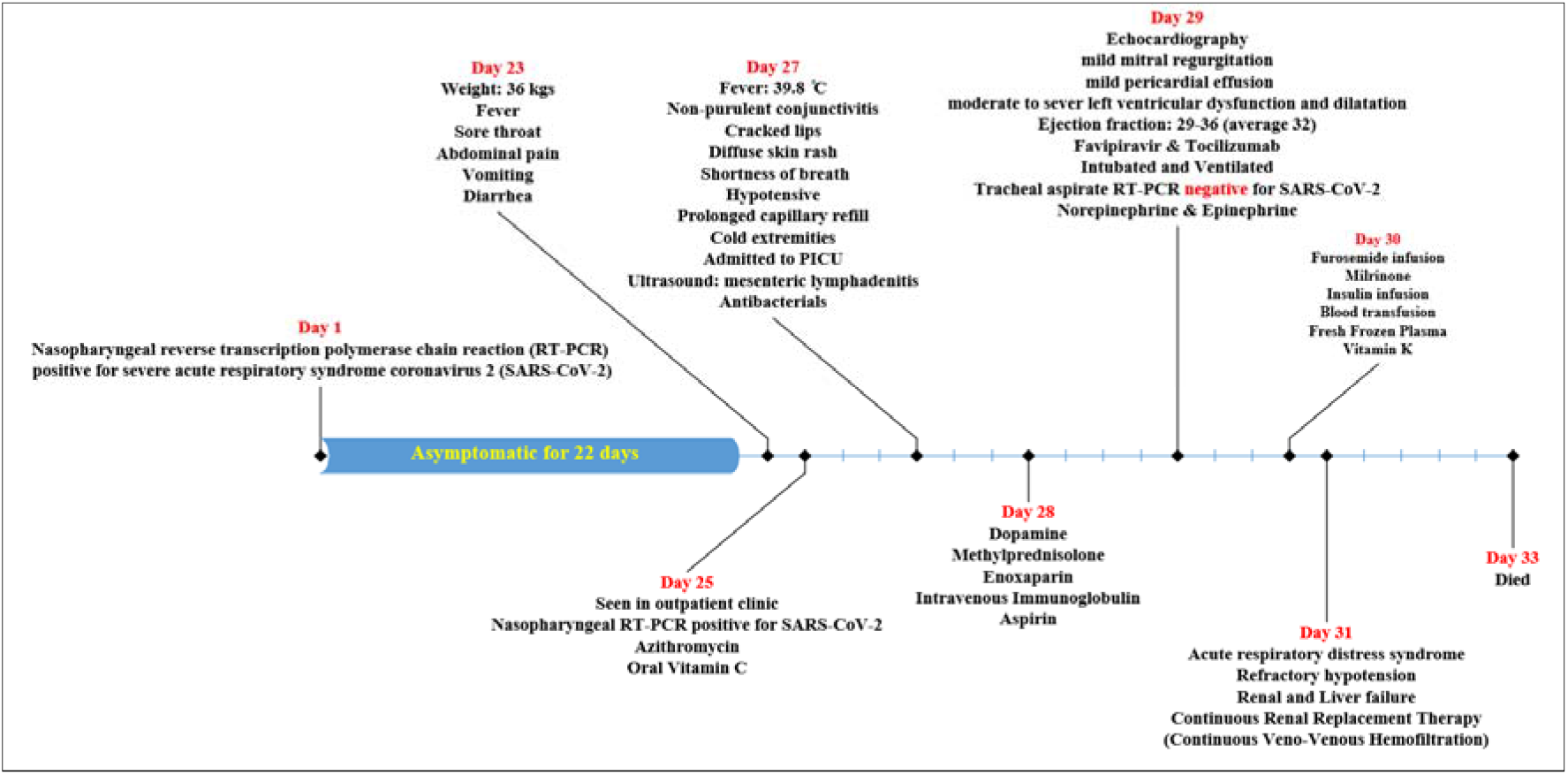
Timeline events for the case.

## Discussion

We report a previously healthy Saudi G6PD deficient girl who died with PMIS after 23 days of being diagnosed as asymptomatic COVID-19.

Our case fulfilled the case definition of PMIS related to COVID-19.^15-17^ Our case aged 10–15 years old, had persistent temperature > 38 □C for ≥ 3 days, acute gastrointestinal problems, clinical features similar to Kawasaki disease, hypotension, shock, myocardial dysfunction, elevated markers of inflammation, evidence of COVID-19, and no plausible alternative diagnoses. The PMIS related to COVID-19 in our case manifested 23 days after confirmed asymptomatic COVID-19. The nasopharyngeal swab of our case continued to be positive for at least 25 days. Her tracheal aspirate sample, which its yield is better than other respiratory swabs,^18^ was negative for COVID-19 after seven days of the PMIS related to COVID-19. We are fairly sure that the two nasopharyngeal swabs were true positive as nasopharyngeal swabs of all her household were positive. These observations confirm that PMIS is delayed inflammatory response after COVID-19.^5, 12^

Our case is the first case of PMIS related to COVID-19 reported in Arabs ethnicity. This syndrome has been reported in Middle Eastern decants living in Europe.^1, 3-5^ However, the Middle East has many different ethnicities, which have not been reported by these studies. The fluorescent spot test (FST) for G6PD screening was positive in our case. This result suggests that our case had a moderate to severe G6PD deficiency as the upper cut-off of our FST is 2.1 Units/ gram Hemoglobin.^19^ This G6PD deficiency makes our case exceptional. African and Afro-Caribbean children overrepresented in reports that have data on race or ethnicity.^1-9^ Although glucose-6-phosphatase dehydrogenase (G6PD) deficiency is common among this population,^14^ the G6PD status of these cases has not been reported in these reports.

G6PD deficiency might contribute to the development of PMIS related to COVID-19 and the grave outcome of our case. G6PD enzyme is present in all body cells, including breast milk.^20, 21^ Redox has a vital part in the modulation of inflammation and immune response.^22^ The G6PD deficiency induces redox imbalance, which exaggerates the inflammatory response.^22^ An ex vivo stud showed that pro-inflammatory interleukin (IL)-8 is upregulated by G6PD deficiency.^22^ The duration of systemic inflammatory response syndrome after major trauma is significantly longer in G6PD-deficient patients compared to their G6PD normal counterpart.^14^ Increased hydrogen peroxide in monocytes of G6PD-deficient major trauma victims has been documented.^14^ Hydrogen peroxide activates pro-inflammatory tumor necrosis factor-alpha (TNF-α).^22^ A recent study suggests that hydrogen peroxide is the first pro-inflammatory messenger.^23^ Monocytes of G6PD deficient major trauma victims have shown to have increased activity and reduced spontaneous apoptosis.^14^ Reduced anti-inflammatory IL-10 level has been evident in monocytes and plasma of type A– G6PD deficient major trauma victims and in Mediterranean G6PD deficient individuals.^14, 24, 25^ It has been shown that alleles of low-producing IL-10, high-producing pro-inflammatory IL-6, and low-producing pro-inflammatory interferon-γ are predominant in G6PD-deficient trauma victims.^26^ Interestingly, it has been demonstrated that ex vivo production of TNF-α, IL-6, and IL-10 in G6PD deficient and G6PD normal infants are the same.^27^ Moreover, the G6PD level is known to decrease with age.^28^ These observations may contribute to why PMIS related to COVID-19 has not been reported in infants yet. G6PD deficiency may be a useful predictor for the progression of the COVID-19. Thus, we urgently need to assess the association between G6PD deficiency and COVID-19 in a large study.

In summary, we report a previously healthy Saudi G6PD Deficient Girl who died with PMIS temporally related to COVID-19. G6PD deficiency may be a useful predictor for the progression of the COVID-19. We believe the association between G6PD deficiency and COVID-19 deserves further study.

## Data Availability

The data are available upon request.

## Conflict of Interest Disclosures (includes financial disclosures)

All authors have no example conflicts of interest to disclose.

## Condolence

We want to express our sincere condolences to the parent and family.

